# A machine learning explanation of the pathogen-immune relationship of SARS-CoV-2 (COVID-19), model to predict immunity, and therapeutic opportunity

**DOI:** 10.1101/2020.07.27.20162867

**Authors:** Eric Luellen

## Abstract

**Importance:** The clinical impacts of this study are it: (1) identified three immunological factors that differentiate asymptomatic, or resistant, COVID-19 patients; (2) identified the levels of those factors that can be used by clinicians to predict who is likely to be asymptomatic or symptomatic; (3) identified a novel COVID-19 therapeutic for further testing; and, (4) ordinally ranked 34 common immunological factors by their importance in predicting disease severity.

**Objectives:** The primary objectives of this study were to learn if machine learning could identify patterns in the pathogen-host immune relationship that differentiate or predict COVID-19 symptom immunity and, if so, which ones and at what levels. The secondary objective was to learn if machine learning could take such differentiators to build a model that could predict COVID-19 immunity with clinical accuracy. The tertiary objective was to learn about the relevance of other immune factors.

**Design:** This was a comparative effectiveness research study on 53 common immunological factors using machine learning on clinical data from 74 similarly-grouped Chinese COVID-19-positive patients, 37 of whom were symptomatic and 37 asymptomatic.

**Setting:** A single-center primary-care hospital in the Wanzhou District of China.

**Participants:** Immunological factors were measured in patients who were diagnosed as SARS-CoV-2 positive by reverse transcriptase-polymerase chain reaction (RT-PCR) in the 14 days before the recordation of the observations. The median age of the 37 asymptomatic patients was 41 years (range 8-75 years), 22 were female, 15 were male. For comparison, 37 RT-PCR test-positive patients were selected and matched to the asymptomatic group by age, comorbidities, and sex.

**Main Outcome:** The primary study outcome was that asymptomatic COVID-19 patients could be identified by three distinct immunological factors and level: stem-cell growth factor-beta (SCGF-β) (> 127637), interleukin-16 (IL-16) (> 45), and macrophage colony-stimulating factor (M-CSF) (> 57). The secondary study outcome was the novel suggestion that stem-cell therapy with SCGF-β may be a new valuable therapeutic for COVID-19.

**Results:** When SCGF-β was included in the machine-learning analysis, a decision-tree and extreme gradient boosting algorithms classified and predicted COVID-19 symptoms immunity with 100% accuracy. When SCGF-β was excluded, a random-forest algorithm classified and predicted COVID-19 asymptomatic and symptomatic cases with 94.8% area under the ROC curve accuracy (95% CI 90.17% to 100%). Thirty-four (34) common immune factors have statistically significant (P-value < .05) associations with COVID-19 symptoms and 19 immune factors appear to have no statistically significant association.

**Conclusion:** People with an SCGF-β level > 127637, or an IL-16 level > 45 and M-CSF level > 57, appear to be predictively immune to COVID-19, 100% and 94.8% (ROC AUC) of the time, respectively. Testing levels of these three immunological factors may be a valuable tool at the point-of-care for managing and preventing outbreaks. Further, stem-cell therapy via SCGF-β and/or M-CSF appear to be promising novel therapeutics for COVID-19.

## Introduction

This section discusses what was known and unknown on this topic, and the resulting hypothesis. This introduction also puts the importance of these findings and the use of machine learning modeling into context.

Asymptomatic patients who are infected with the SARS-CoV-2 virus have neither clinical symptoms nor abnormal chest imaging. However, asymptomatic patients have the same infectivity as infected patients with symptoms [1]. Moreover, adult asymptomatic patients have been found to have the same viral loads as symptomatic patients [2]. Studies have shown that age appears to influence whether an infected person is susceptible to illness. Those under the age of 20 years have approximately half the morbidity probability as those over the age of 20 [3]. This improbability of becoming ill from the SARS-CoV-2 virus is especially interesting because young children have been found to have 10 to 100 times the viral load as older children and adults, and disproportionately remain asymptomatic [4].

SCGF-β has been associated with H7N9 (Asian lineage avian influenza A subtype) and disassociated with H5N1 (highly pathogenic avian influenza (HPAI) [5][6].

Elevated SCGF-β has also been associated with the specific disease states of hepatocellular cancer, Chagas’ disease, cardiomyopathy, inflammation and insulin resistance, and unstable carotid plaques [7] [8] [9][10]. Interleukin 16, the second most important variable in predicting SARS-CoV-2 immunity or resistance here, has been strongly associated with asthma [11].

Prior studies on the biomarkers associated with SARS-CoV-2 immune response and morbidity include interferon-gamma (IFN-γ), interferon-beta (IFN-β), and interleukin-8 (IL-8) [12]. Other previous research on immune parameters associated with SARS-CoV-2 severity and prognosis have involved interleukin one beta (IL-1β) and interleukin six (IL-6). However, others found reduced immunoglobin G levels in asymptomatic patients [13][14]. The general finding in prior research regarding the pathogen-immune relationship with SARS-CoV-2 is that symptomatic patients have considerably more inflammation and cytokine storm activity than asymptomatic patients [14].

What has been unknown for SARS-CoV-2 are three questions to which the answers are suggested in this study. First, which immunological variables are statistically significant, and how important is each in predicting asymptomatic status? Second, which of those variables, if any, have a strong negative correlation, or relationship, with disease severity (i.e., asymptomatic patients’ levels are significantly higher than symptomatic patients)? And third, is there an algorithmic or formulaic model of prognostic biomarkers that can accurately predict morbidity – who will be asymptomatic if infected, and who is at risk of more severe symptoms and disease progression – and why?

## Methods

This study was based on secondary data published as a supplement in Nature Medicine in June 2020. Therein immunological factors were measured in 74 patients in the Wanzhou District of China. They were diagnosed as SARS-CoV-2 positive by reverse transcriptase-polymerase chain reaction (RT-PCR) in the 14 days before the recordation of the observations. The median age of the 37 asymptomatic patients was 41 years (range 8-75 years), 22 were female, 15 were male. For comparison, 37 RT-PCR test-positive patients were selected and matched to the asymptomatic group by age, comorbidities, and sex [14].

In this study, five algorithms, or types, of machine learning – a kind of artificial intelligence employing robust brute-force statistical calculations – were applied to a data set of 74 observations of 34 immunological factors to attempt to do three things: (1) develop a model to accurately predict the classification of which patients will be asymptomatic or symptomatic to SARS-CoV-2; (2) determine the relative importance of each immunological factor; and, (3) determine if there is any level of a subset of immunological factors that can accurately predict which patients are likely to be immune or resistant to SARS-CoV-2.

Minitab 19 (version 19.2020.1, Minitab LLC) was used to calculate means, 95% confidence intervals, P-values, and two-sample T-tests of statistical significance. Correlation coefficients were also computed using Minitab via Spearman rho because the data was distributed nonparametrically. A second classification and regression tree (CART) algorithm were also applied in Minitab to cross-validate decision tree results from R in Rattle. Minitab’s CART methodology was initially described by Stanford University and the University of California Berkeley researchers in 1984 [15].

The Rattle library (version 5.3.0, Togaware) in the statistical programming language R (version 3.6.3, CRAN) was used to apply five machine learning algorithms – a decision tree, extreme gradient boosting (XGBoost), linear logistic model (LLM), random forest, and support vector machine (SVM) – to learn which model, if any, could predict asymptomatic status, how accurately, and how. Rattle randomly partitioned the data to select and train on 80% (n=59), validate on 10% (7), and test on 10% (7) of observations. Two evaluation methods were used: (1) plots of linear fits of the predicted versus observed categorization; and, (2) a pseudo-R^2^ measure calculated as the square root of the correlation between the predicted and observed values. Pseudo-R^2^ measure results were evaluated twice, each using for evaluation data that were held back by being randomly selected during partitioning and averaging the two accuracy findings for the final results.

Rattle’s rpart decision tree was also used to identify if any levels of one or more immunological factors that could accurately diagnose someone was asymptomatic (i.e., via rules). The decision tree results reported here used 20 and 12 as the minimum number of observations necessary in nodes before the split (i.e., minimum split). The trees used 7 and 4 as the minimum number of observations in a leaf node (i.e., minimum bucket).

The random forest analysis in Rattle began by running a series of differently sized random forest algorithms, ranging from 50 to 500 decision trees, to learn the optimum number of trees to minimize error. Each random forest consisted of a minimum of six variables, which was closest to the square root of the number of statistically significant variables, 34. The lowest error rate was approximately 200 decision trees, which was applied, using four variables at a time, which was the closest whole number to the square root of the number of predictors.

The five machine learning models and CART classification trees were run, including, and excluding, SCGF-β to identify if there were alternative prognostic biomarkers and levels in the immune profile that could accurately classify and predict SARS-CoV-2 immunity.

## Results

Thirty-three (34) of the 53 immunological factors (64.2%) were indicated as statistically significant by P-values less than .05 from a Spearman rho correlation. Of those 34 factors, 31 were statistically significant with P-values less than .01. Conversely, 35.9% of the 53 immune factors had no statistically significant association with whether a patient was asymptomatic or symptomatic to SARS-CoV-2.

The 22 factors positively correlated with being symptomatic ranged from a minimum coefficient of .205 (MCP-3) to a maximum of .781 (TRAIL). The 11 factors negatively associated with being symptomatic ranged from a minimum of -.866 (SCGF-β) to a maximum of -.276 (IFNα2) (see Table 1).

**Table 1:** Immunological factors associated with SARS-CoV-2 morbidity ranked by Spearman correlation coefficients with 95% confidence intervals and P-values (statistically insignificant and corresponding P-values in gray text; negative correlations highlighted in gray at bottom of the table)

Figures
| Immunological Factor | Abbreviation | Pairwise Spearman Correlation to Asymptomatic (0) or Symptomatic (1) Status | 95% CI | P-Value (< .05 target) |
| --- | --- | --- | --- | --- |
| TNF-related apoptosis-inducing ligand | TRAIL | 0.781 | (0.654, 0.865) | < .000 |
| Growth-regulated oncogene alpha | GRO-α | 0.75 | (0.611, 0.845) | < .000 |
| Macrophage colony stimulating factor | M-CSF | 0.748 | (0.608, 0.843) | < .000 |
| Interleukin-6 | IL-6 | 0.705 | (0.549, 0.813) | < .000 |
| Granulocyte colony-stimulating factor | G-CSF | 0.697 | (0.539, 0.808) | < .000 |
| Interleukin-2 | IL-2 | 0.667 | (0.499, 0.787) | < .000 |
| Nerve growth factor beta | NGF-β | 0.651 | (0.479, 0.775) | < .000 |
| Interleukin-10 | IL-10 | 0.614 | (0.431, 0.748) | < .000 |
| Monocyte chemoattractant protein-1 | MCP-1 | 0.594 | (0.407, 0.733) | < .000 |
| Stem cell factor | SCF | 0.586 | (0.397, 0.728) | < .000 |
| Interleukin-15 | IL-15 | 0.527 | (0.325, 0.683) | < .000 |
| Interleukin-8 | IL-8 | 0.514 | (0.311, 0.673) | < .000 |
| Interferon gamma | IFN-γ | 0.464 | (0.252, .633) | < .000 |
| Interleukin-7 | IL-7 | 0.454 | (0.240, 0.625) | < .000 |
| Interferon gamma inducible protein-10 | INF-γ-IP-10 | 0.451 | (0.237, 0.623) | < .000 |
| Interleukin-18 | IL-18 | 0.438 | (0.223, 0.613) | < .000 |
| Platelet derived growth factor-BB | PDGF-BB | 0.436 | (0.220, 0.611) | < .000 |
| Interleukin-2 receptor alpha | IL-2Rα | 0.388 | (0.166, 0.572) | 0.001 |
| Immunoglobulin G (convalescing) | IgG Convalesce | 0.366 | (0.143, 0.544) | 0.001 |
| Monokine induced by gamma | MIG | 0.364 | (0.140, 0.552) | 0.001 |
| Immunoglobulin G (acute) | IgG-Acute | 0.33 | (0.103, 0.524) | 0.004 |
| Macrophage migration inhibitory factor | MIF | 0.237 | (0.006, 0.444) | 0.042 |
| Monocyte chemotactic protein-3 | MCP-3 | 0.205 | (-.027, 0.416) | 0.079 |
| Vascular endothelial growth factor | VEGF | 0.184 | (-0.048, .397) | 0.117 |
| N gene | N | 0.18 | (-0.053, 0.394) | 0.126 |
| Interleukin-3 | IL-3 | 0.163 | (-0.070, 0.379) | 0.166 |
| Interleukin-12-p40 | IL-12(p40) | 0.151 | (-0.082, 0.368) | 0.199 |
| Interleukin-9 | IL-9 | 0.149 | (-0.084, 0.366) | 0.206 |
| Interleukin-1 beta | IL-1β | 0.125 | (-0.107, 0.345) | 0.287 |
| Days shed virions | Days shed | 0.122 | (-0.110, 0.342) | 0.3 |
| Stromal cell-derived factor-1 alpha | SDF-1α | 0.098 | (-0.134, 0.320) | 0.406 |
| Interleukin-12-p70 | IL-12(p70) | 0.083 | (-0.149, 0.306) | 0.484 |
| Interleukin-17 | IL-17 | 0.067 | (-0.164, 0.291) | 0.57 |
| Interleukin-4 | IL-4 | 0.02 | (-0.210, 0.247) | 0.868 |
| Interleukin-13 | IL-13 | -0.022 | (-0.249, 0.208) | 0.856 |
| Fibroblast growth factor | FGF | -0.078 | (-0.302, 0.153) | 0.506 |
| Regulated upon activation, normal T cell expressed and secreted | RANTES | -0.085 | (-0.308, 0.146) | 0.469 |
| Macrophage inflammatory protein-1 beta | MIP-1β | -0.109 | (-0.330, 0.123) | 0.353 |
| ORF1ab gene | ORF1ab | -0.113 | (-0.334, 0.119) | 0.337 |
| Macrophage inflammatory protein-1 alpha | MIP-1α | -0.138 | (-0.356, 0.095) | 0.241 |
| Tumor necrosis factor-alpha | TNF-α | -0.168 | (-0.383, 0.065) | 0.153 |
| Tumor necrosis factor-beta | TNF-β | -0.197 | (-0.409, 0.035) | 0.093 |
| Interferon alpha-2 | IFNα2 | -0.276 | (-0.478, -0.046) | 0.017 |
| Leukemia inhibitory factor | LIF | -0.312 | (-0.509, -0.84) | 0.007 |
| Interleukin-5 | IL-5 | -0.316 | (-0.512, -0.089) | 0.006 |
| Interleukin-1 alpha | IL-1α | -0.332 | (-0.526, -0.106) | 0.004 |
| Granulocyte-macrophage colony-stimulating factor | GM-CSF | -0.359 | (-0.548, -0.134) | 0.002 |
| Interleukin-1 receptor alpha | IL-1Rα | -0.359 | (-0.548, -0.135) | 0.002 |
| Eotaxin | Eotaxin | -0.39 | (-0.576, -0.169) | 0.001 |
| Cutaneous T cell-attracting chemokine | CTACK | -0.456 | (-0.627, -0.243) | < .000 |
| Hepatocyte growth factor | HGF | -0.594 | (-0.733, -0.407) | < .000 |
| Interleukin-16 | IL-16 | -0.827 | (-0.895, -0.721) | < .000 |
| Stem cell growth factor beta | SCGF-β | -0.866 | (-0.92, -0.78) | < .000 |

When SCGF-β was included in the machine-learning analysis, two algorithms predicted and classified SARS-CoV-2 immunity or resistance by being asymptomatic with 100% accuracy: a decision tree, and XGBoost. When SCGF-β was excluded, a random-forest algorithm predicted and classified SARS-CoV-2 asymptomatic and symptomatic cases with 94.8% area under the ROC curve accuracy (95% CI 90.17% to 100%) (see Table 2).

**Table 2:** Comparative accuracy of six machine learning algorithms in predicting SARS-CoV-2 asymptomatic status from immunological factors.

| Machine Learning Model | Pseudo R2 (10% evaluation holdback sample 1) | Pseudo R2 (10% evaluation holdback sample 2) | Average Pseudo R2 |
| --- | --- | --- | --- |
| With SCGF-β | | | |
| Decision tree | 100.00% | 100.00% | 100.00% |
| XGBoost | 100.00% | 100.00% | 100.00% |
| GLM (logistic) | 100.00% | 98.89% | 99.45% |
| Random forest | 99.46% | 94.83% | 97.15% |
| SVM | 78.81% | 96.99% | 87.90% |
| Without SCGF-β | | | |
| Random forest | 97.68% | 91.91% | 94.80% |
| GLM (logistic) | 100.00% | 85.96% | 92.98% |
| SVM | 77.76% | 89.69% | 83.73% |
| XGBoost | 99.42% | 54.27% | 76.85% |
| Decision tree | 100.00% | 2.22% | 51.11% |

Notably, both the rpart decision trees and CART classification trees independently identified three prognostic biomarkers at specific levels that could classify asymptomatic and symptomatic cases with 95-100% accuracy. When SCGF-β was included, all asymptomatic cases had levels > 127656.8, while all symptomatic cases had levels < 127656.8 (see Figure 1). When SCGF-β was excluded, as a type of contingency analysis to understand prognostic biomarker levels in other factors better, IL-16 accurately classified asymptomatic cases > 44.59 and symptomatic cases < 44.59 in 90.4% of the cases. In the remaining 9.6% of cases where IL-16 > 44.59, all of them had M-CSF > 57.13 (see Figure 2).

**Figure 1:**
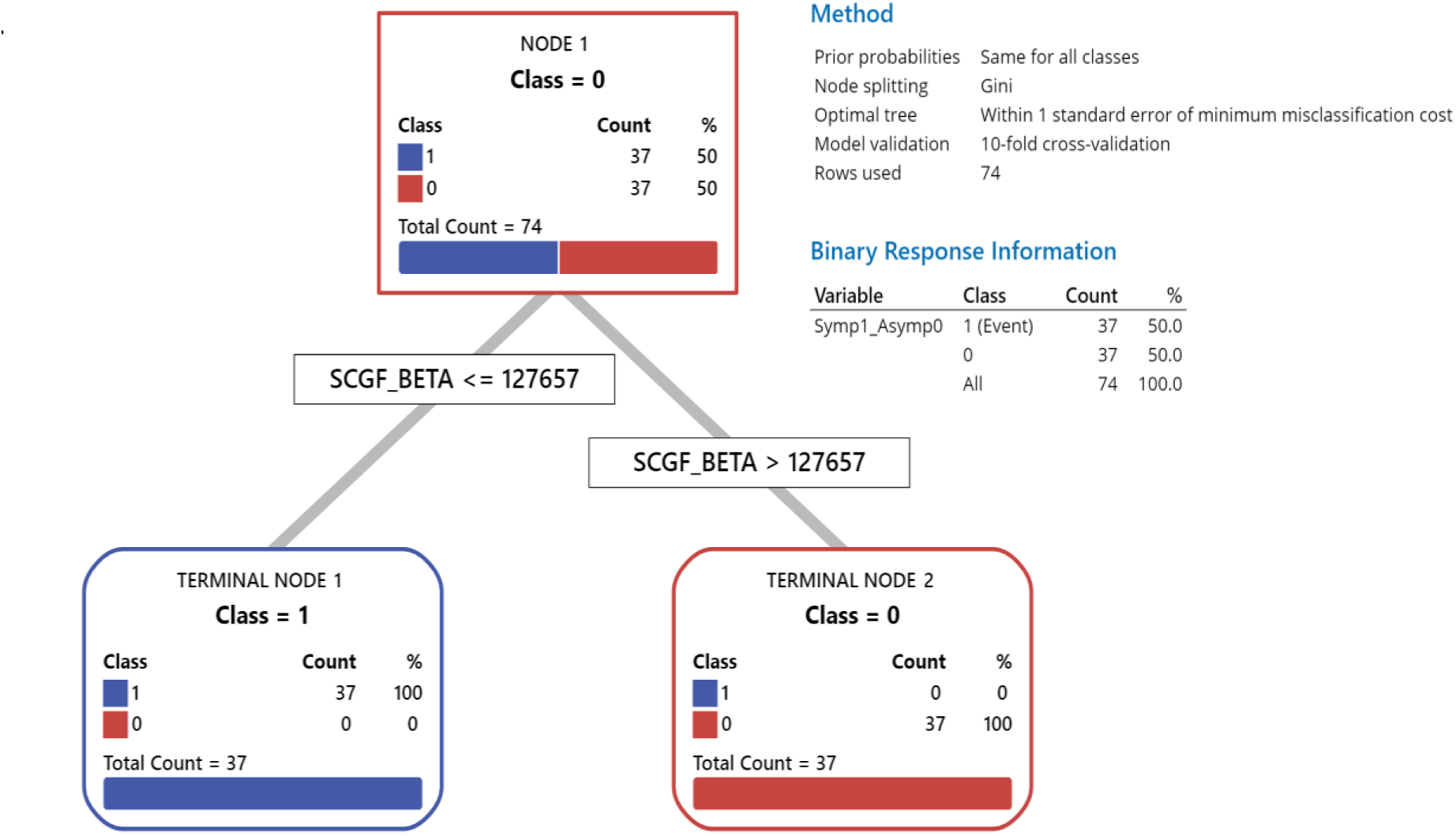
CART classification tree of role of SCGF-β in predicting SARS-CoV-2 morbidity.

**Figure 2:**
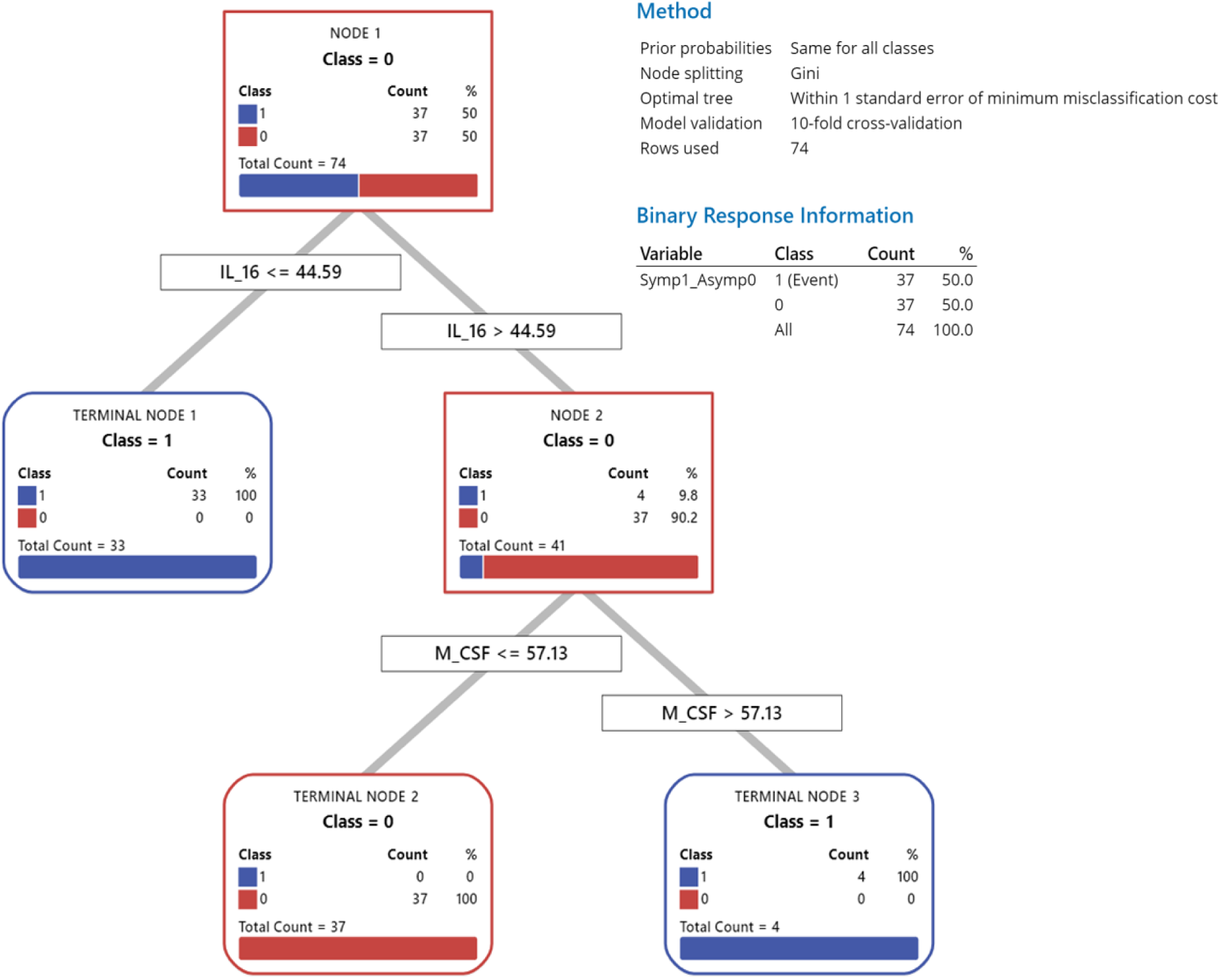
CART classification tree of rule of IL-16 and M-CSF in predicting SARS-CoV-2 morbidity in the absence of SCGF-β.

Two-sample T-tests for the four factors with the highest positive and negative correlation coefficients, interquartile ranges, outliers, and levels between asymptomatic and symptomatic patients that were statistically significant were computed to ordinally rank factors by their correlation coefficients (see Figure 3).

**Figure 3:**
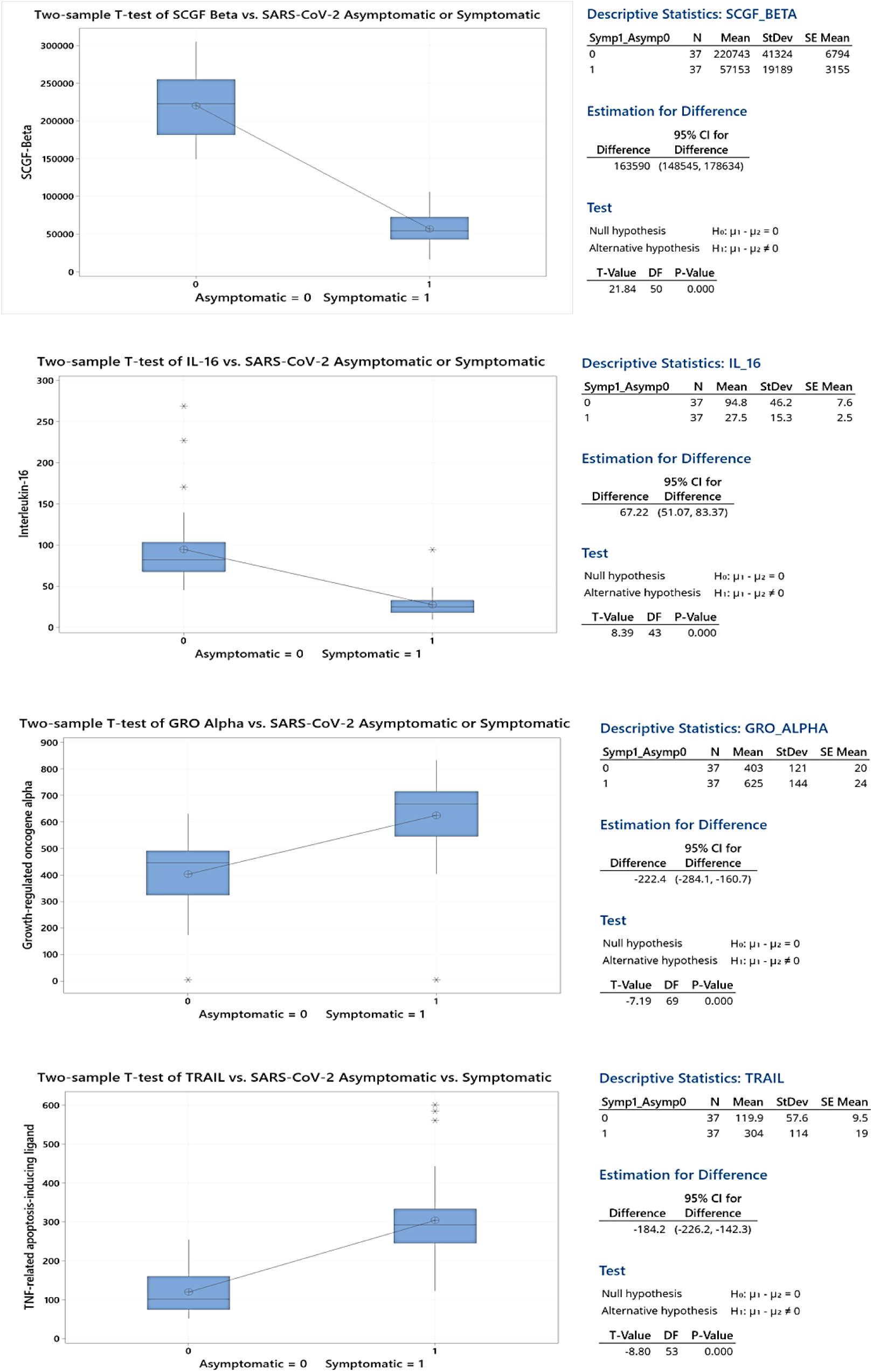
Two-sample T-tests of statistical significance of difference in means of four leading prognostic biomarkers for asymptomatic or symptomatic SARS-CoV-2.

A random forest analysis of the most important variables to accurately classify and predict SARS-CoV-2 patients by binary morbidity ordinally ranked the 33 statistically significant factors. Unsurprisingly, SCGF-β, and IL-16, followed by GRO-α and TRAIL, respectively, were the most critical factors in predicting morbidity (see Figure 4).

**Figure 4:**
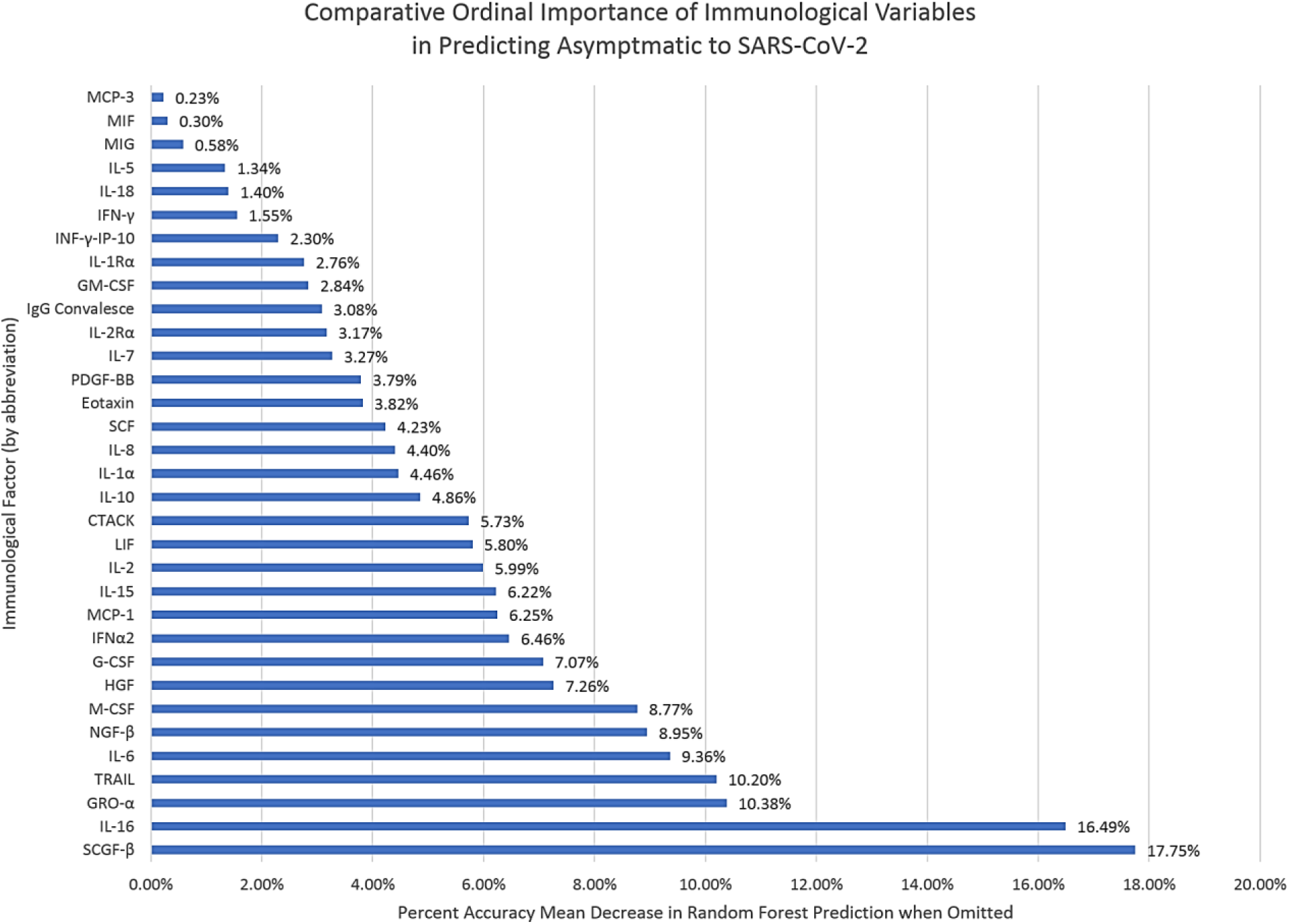
Relative importance of immunological variables from random forest analysis in predicting SARS-CoV-2 morbidity.

Finally, the results suggest that IL-1β, 3, 4, 9, 12, 13, 17, and RANTES are of low importance, or comparative irrelevance, in the pathogen-immune relationship and, that SCGF-β, IL-16, HGF, INFNα2, LIF, CTACK, IL-1α, Eotaxin, GM-CSF, IL-1Rα, and IL-5 are critical in models to predict and classify asymptomatic or symptomatic SARS-CoV-2 cases accurately.

## Discussion

While it has been speculated that stem cells may play a role in SARS-CoV-2 and other zoonoses’ resistance, prior research has focused on different stem cell involvement than stem-cell growth factorbeta [16][17][18]. Previous research has also established that stem cells can inhibit viral growth by expressing interferon-gamma stimulated genes (ISGs) and have been particularly effective against influenza A H5N1 virus and resulting lung injuries [19][20]. Stem cell therapy (SCT) has been hypothesized as a treatment for SARS-CoV-2; however, there is no record in the literature specific as to which factors may influence SARS-CoV-2 infections, favorably or unfavorably, or to what degree until now [21].

Researchers have recently found that symptomatic patients generally have a more robust immune response to SARS-CoV-2 infection, culminating in cytokine storms in the worst cases. Conversely, asymptomatic patients have been found to have a weaker immune response [14]. Because infections are causal to immune response, of particular interest in this study were the most impactful immune-related variables that negatively correlated with asymptomatic status (i.e., variables that were greater for asymptomatic patients than symptomatic patients), which are highlighted in gray in Table 1.

This work’s overarching importance is the identification of immunological factors for diagnoses, treatments, and pre-clinical prophylactic immune-based approaches to SARS-CoV-2 in the first seven months of a pandemic that experts now opine will last decades [22]. Immunostimulant approaches are especially valuable because, unlike antivirals and vaccines, they may be given later in the course of the disease to optimize outcomes [21].

The primary importance of this work is machine learning algorithmic models that can predict with high accuracy, whether someone, once infected, will be asymptomatic or symptomatic from SARS-CoV-2. This knowledge gives clinicians new tools to identify populations in advance who appear to be at higher risk of danger from the virus. Such devices, especially once reproduced in a more extensive study, may also inform policy decisions as to who needs to shelter-in-place. Finally, because of the scale of this pandemic and practical constraints as to how many vaccination doses can be manufactured how quickly, such tools may become valuable in prioritizing vaccine administration to those in greatest need because they are at the higher biological and immunological risk.

This work’s secondary importance is a description of the cytokine and chemokine profile that is associated with asymptomatic or symptomatic SARS-CoV-2 infections. It enables a better understanding of the pathogen-immune relationship. These profiles provide insights into the biological pathways critical for SARS-CoV-2 progression.

As one example, stem cell factors secrete multiple factors that regulate immune cells and modulate them to restore tissue homeostasis. These results suggest that higher levels of SCF-β may better control immune responses to prevent the more robust reactions universally associated so far with highly symptomatic patients and, further, prevent high morbidity and mortality cytokine storms. A better understanding of the pathogen-immune relationship may enable researchers to prevent and treat SARS-CoV-2 patients more effectively with therapeutics currently untested and unused. This knowledge may also extend to similar zoonotic coronaviruses in the future.

The tertiary importance of this work is identifying three immune factors and precise levels that appear to be prognostic biomarkers as to whether someone, once infected with the SARS-CoV-2 virus, will be immune or resistant, as demonstrated by being asymptomatic, or not. These insights also suggest new candidates for therapeutic research focused on the relatively newly identified and ill-understood SCGF-β and its role in the immunological process.

The quaternary importance of this work is further proof that machine-learning methods can accurately and quickly identify critical elements of disease dynamics that accelerate understanding and improve outcomes during pandemics. Moreover, it is an example of how a ‘dry’ data science laboratory can link to clinical or ‘wet’ laboratory science for real-world applications.

This study has several limitations. First, it is unknown from the dataset how many days passed between exposure to the virus and immunological testing, or whether it was universally the same number of days. Second, because immune profiles are temporally sensitive, ideally, several tests would have been taken over several days, which did not occur [23]. Third, immunological signaling and processing are multifactorial and complex. Therefore, it is unclear why SCGF-Beta levels are categorically high in asymptomatic patients and low in symptomatic patients, or whether they are causal to SARS-CoV-2 response. Fourth, combinatorial and sequential analysis of these immunological elements may be an important future research area to optimize therapeutic research outcomes. Fifth, at least one study in a leading journal, *Lancet*, found that Chinese SARS-CoV-2 case data may have been misreported by as much as 400% [24]. That study, and much higher case and fatalities numbers in over 200 countries, have created distrust and skepticism of SARS-CoV-2-related data originating in China.

Future research could ameliorate these limitations and focus on a more extensive study group to attempt to reproduce the results. Moreover, a prospective case-control study of patients with decreased SCFG-β levels and supplementation was protective against SARS-CoV-2 severity and symptoms.

## Conclusion

One implication of these findings is that if we can predict the 80% of society who may be immune or resistant to SARS-CoV-2, or asymptomatic, it may profoundly impact public health intervention decisions as to who needs to be protected and how much. If, for example, 80% of the shelter-in-place orders and the resultant dramatic reduction in economic and social activity could have been prevented by accurately predicting who is at low risk of infection, the economic benefits alone may have been valued in US$ trillions. The second implication of these findings is evidence that elevated levels of SCGF-β, IL-16, and M-CSF may have a causal relationship with SARS-CoV-2 immunity or resistance may have utility as diagnostic determinants to (a) inform public health policy decisions to prioritize and reduce shelter-in-place orders to minimize economic and social impacts; (b) advance therapeutic research; and, (c) prioritize vaccine distribution to benefit those with the greatest need and risks first.

## Data Availability

The data is in the public domain.

## Acknowledgment

The author wishes to thank Dr. Luka Fajs for his helpful comments on drafts of this article.Figures

## Notes

### Competing Interest Statement

The authors have declared no competing interest.

### Funding Statement

No external funding was received for this study.

### Summary of Updates

This third version had three modifications: (1) the title was clarified; (2) a structured abstract was added; and, (3) references were converted from APA parenthetical citations to numbered-by-appearance journal formatting.

